# A systematic review and modelling insights of factors impacting measles vaccine effectiveness

**DOI:** 10.1101/2024.08.29.24312705

**Authors:** Samiran Ghosh, Divya Kappara, Nabanita Majumder, Suchita Nath-Sain, Arup Deb Roy, Siuli Mukhopadhyay

## Abstract

**Introduction:** Outbreaks of measles have been frequently reported despite the availability of an effective vaccine. In this systematic review we examine the potential factors that could impact the effectiveness of the measles vaccine (MV) in children.

**Methods:** We conducted a literature search using PubMed and Google Scholar for primary articles published between January 2014 and July 2024. Articles reporting the effectiveness or immunogenicity of MV in children aged 0-15 years were included. Additionally, we use regression analysis on data available from few cohort studies in India and epidemiological modelling simulations to assess the effect of factors on vaccine effectiveness (VE).

**Results:** Overall, 34 primary articles involving 151888 children who had received the MV were included in the analysis. Key factors that may affect VE were malnutrition, genetic variants, chemotherapy and gender. Through statistical modelling we identified an inverse relationship between malnutrition and measles VE and estimated the possible percentage decrease in VE due to malnutrition across different states of India. Additionally, simulations from a Susceptible-Infected type model showed the effect of varying VE on the modelling outcomes and measles elimination targets.

**Conclusions:** We identified a few key factors including malnutrition, genetic variants, chemotherapy and gender that impact measles VE. Therefore, in addition to maintaining WHO recommended vaccine coverages, addressing the problems related to VE is crucial for achieving measles elimination targets.

## INTRODUCTION

Measles is a highly contagious, vaccine-preventable viral disease transmitted by respiratory droplets, small particle aerosols, and close contact [1]. Each infection can cause 12-18 secondary cases among the at-risk population. According to recent estimates from the World Health Organization (WHO) and the U.S. Centers for Disease Control and Prevention (CDC), global measles cases reached approximately 10.3 million in 2023, marking a 20% rise compared to 2022 [2]. Due to gaps in vaccination coverage worldwide, 57 countries across the WHO African, Eastern Mediterranean, European, Southeast Asian, and Western Pacific regions witnessed a substantial upsurge in measles cases [2].

Vaccination against measles at the community level has been the most effective way to prevent the disease [3]. The World Health Organization (WHO) currently recommends administering the first dose of the vaccine at 9 months in areas where measles is common and at 12-15 months elsewhere. A second dose is recommended, usually at 15-18 months. Serologic evaluations have shown that the measles vaccine (MV) elicits immune responses in the majority of the susceptible population with median vaccine effectiveness (VE) of 84% for one dose and 94.1% for two doses [4].

Although an effective vaccine is available, measles virus remains an important cause of worldwide mortality and morbidity accounting for 107,500 deaths in 2023 [3].

While measles vaccination has prevented ∼60 million deaths between 2000-2023, it is still a common disease in many parts of the world with yearly outbreaks. The majority of these deaths occur among children under the age of five, particularly in regions where vaccination coverage is low or inconsistent [5]. In 2023, measles cases in Southeast Asian countries surged to 40.9 per million from 16.2 per million population in 2013 even though 91% of children received one dose and 85% received two doses of MV [6]. In India particularly, which report much higher measles vaccine (MV) coverage (93.35% for first dose, and 89.84% for two doses [6]) than the global or the South-East Asian coverages, several recent outbreaks have threatened the elimination efforts in the country. These high disease counts and repetitive outbreaks raise questions about the actual effectiveness of the MV [7, 8]. It is worrisome that such an effective [4] and high coverages were unable reduce the frequency of the measles outbreaks. As noted in the literature, the effectiveness of the MV has a crucial role to play in the prevention of outbreaks and successful control of the spread of the measles virus. Thus, any possibility of lowered vaccine effectiveness leading to these frequent outbreaks raises concerns regarding any country’s progress toward elimination of the disease. Such observations motivate us to search and evaluate the effect of any underlying factors in the susceptible populations of such countries which may be affecting and lowering the VE, thus making them the population more vulnerable to the disease.

Vaccine effectiveness (VE), i.e the ability of vaccines to provide protection in real-world conditions [9], has been noted to vary in different populations, with lower estimates particularly in African [10] and South East Asian countries. Individual variation in the immune response to vaccination for several diseases including measles, have been reported widely in the literature [11–15]. Zimmerman et al [15] reported that an estimated 19 million measles vaccinated children have been left unprotected due to ineffectiveness of MV.

Given the paucity of research in this area, in this systematic review, our aim is to identify the important factors, which may be responsible for lowering the VE of MV in children. Understanding and identifying these factors is essential for addressing the challenges posed by variations in vaccine performance across different populations. By identifying the socio-demographic and other contributory factors that may reduce the VE of MV, this review seeks to provide actionable insights for improving public health interventions. The findings of this review are expected to aid policymakers in tailoring disease control and elimination strategies, ensuring they account for regions or populations with suboptimal vaccine effectiveness. Additionally, we assess the effect of a key factor of VE using statistical tools and study the impact of varying VE on epidemiological model predictions through simulations. Variations in VE is shown to substantially effect model-generated estimates of disease burden.

## METHODS

We conducted a systematic review according to the Preferred Reporting Items for Systematic Reviews and Meta-Analyses (PRISMA) statement [16].

### Search strategy

We searched PubMed and Google Scholar for primary articles published between January 2014 and July 2024. using Medical Subject Headings (MeSH) terminology for our population of interest including infants, preterm infants, premature birth, premature infants, children or adolescents; interventions including measles-mumps-rubella vaccine, MMR vaccine, pluserix, virivac, trimovax, triviraten berna, or priorix; and outcomes including VE, vaccine immunogenicity, or antibody response (**Table 1**). While our intention was to study VE in real-world conditions, this term has been used interchangeably with ‘vaccine efficacy’ or the effect of vaccines in controlled clinical trial settings [9]. Therefore, articles reporting both these terms were included.

**Table 1.** Population, intervention, comparison, and outcome (PICO) framework

| **PICO Framework** |  |
| --- | --- |
| **P (Population)** | Children aged 0–15 years who have received the measles vaccine (MV) |
| **I (Intervention)** | Measles vaccine |
| **C (Comparison)** | Not applicable |
| **O (Outcome)** | The primary outcome of this systematic review is to identify the important factors that may be responsible for lowering the vaccine effectiveness (VE) of MV in children. Additionally, to assess the effect of a key factor of VE using statistical tools and study the impact of varying VE on epidemiological model predictions through simulations |

Vaccine immunogenicity refers to the ability of vaccines to induce a measurable immune response [17]. The literature search was limited to articles reported in English. Additionally, articles were identified from websites as well as from references cited in review articles.

### Inclusion and exclusion criteria

Primary articles reporting the effectiveness or immunogenicity of MV in children aged 0-15 years were included. Narrative or systematic reviews, meta-analysis, book chapters, letters, as well as articles reporting other disease areas or not meeting the inclusion criteria were excluded.

### Selection and data extraction

Articles retrieved from the databases were imported to EndNote 21. Duplicate articles were removed. The articles were independently screened by two authors based on titles and abstracts. Relevant data were extracted to a spreadsheet from the selected articles by two independent authors. Study-level information was recorded in the spreadsheet, which included author names, year of publication, study period, study design, setting, sample size, age group, and factors impacting measles VE. Discrepancies regarding inclusion of individual articles were resolved by discussion with a third author.

### Assessment of risk of bias

The risk of assessment was conducted by three independent authors using the Joanna Briggs Institute (JBI) Critical Appraisal tool [18] (Supplementary tables I-IV). Bias was assessed at the study, outcome, and result level. The tool has a specific number of questions for each study type (case-control, cohort, cross-sectional and randomised control trials). Each question is assessed by scoring (yes=1), (no=0) and (unclear or not applicable=0). Based on the total score, each study was categorised as high (20-49%), moderate (50-79%) or low (80-100%) risk. Risk of bias was assessed by three authors and discrepancies were resolved by consensus.

### Statistical analysis

We utilised linear regression to assess the statistical relationships and a Susceptible-Infected (SI) model as described previously [19, 20], to simulate the scenario analysis. Data were processed and analysed using R for statistical modelling and MATLAB for the SI simulations. The spatial visualisations were created in R using publicly available geographic datasets from https://onlinemaps.surveyofindia.gov.in.

## RESULTS

### Study selection and characteristics

A total of 514 articles were retrieved from databases (PubMed, 334; Google Scholar, 180), of which 11 duplicates were excluded and 503 were screened based on title/abstract (**Figure 1**). Of these, 460 articles were excluded as they were not primary articles or did not meet the prespecified inclusion criteria. The full-text for 1 article was unavailable leaving 42 articles for review. An additional 37 articles were identified from websites and cross-references of which 12 were considered for full-text review. After full-text review of a total of 54 articles, 20 articles were excluded as they were not primary articles or did not meet the inclusion criteria. Overall, 34 studies were included in the analysis.

**Figure 1.**
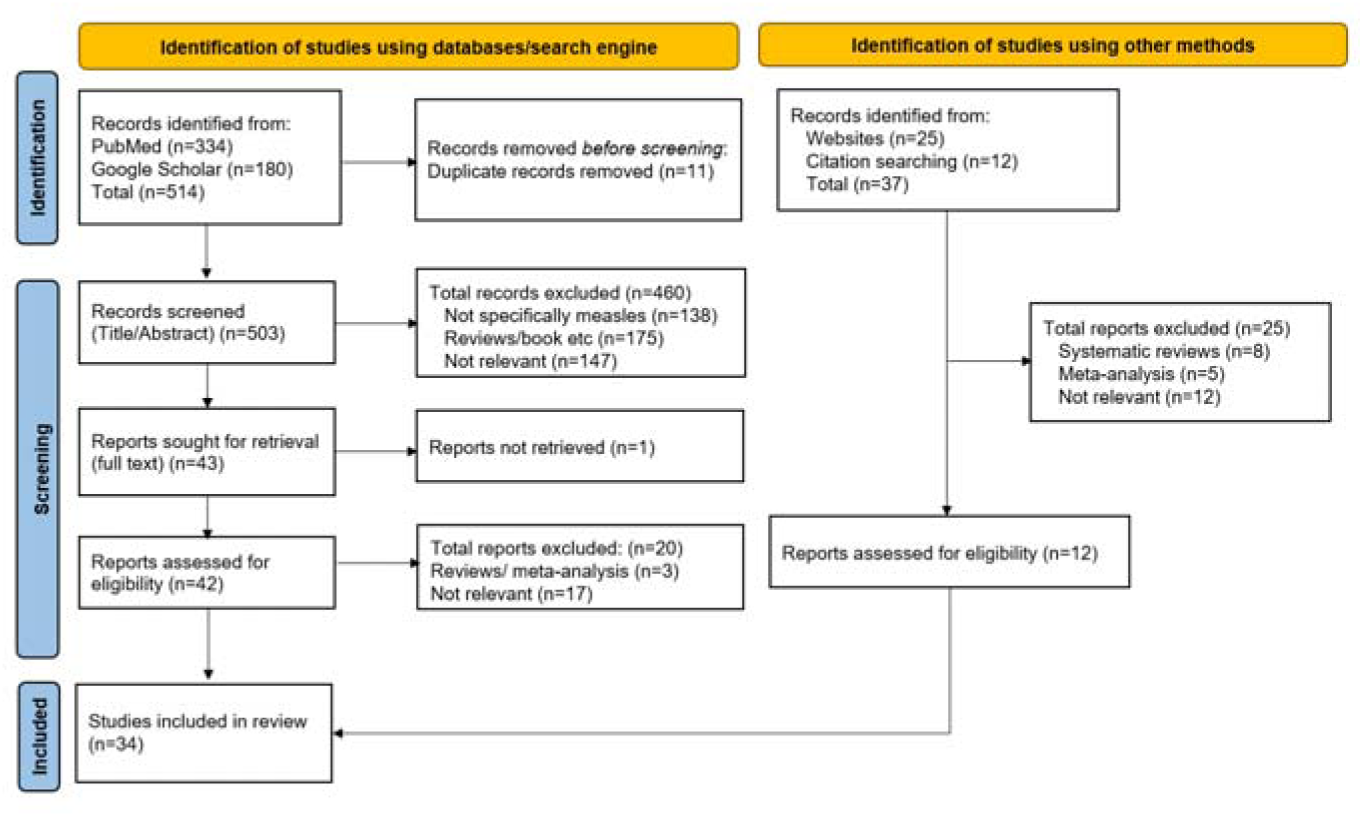
PRISMA flow diagram

The studies were conducted in several countries including Australia, Brazil, Czech Republic, Democratic Republic of Congo, Guinea-Bissau, Italy, Kuwait, and United States. A total of 8 studies were conducted in Asia of which 1 was from India (**Table 2**). Most were cohort studies (n=15), followed by randomised controlled trials (RCTs; n=8), cross-sectional (n=6) and case-control studies (n=5). Community (number of studies=17) and hospital-based studies (n=7) were most common. The studies included 151888 children comprising 7329 infants (0-12 months).

**Table 2.** Study characteristics of the included articles

| Author, year | Country | Study period | Study design | Study setting | Sample size | Age group | Factors affecting the effectiveness of measles vaccine |
| --- | --- | --- | --- | --- | --- | --- | --- |
| Bati et al, 2021 | Ethiopia | 2014 | Case-control | Community | 204 | 6-59 months | Age at first dose and Cold chain |
| Carryn et al, 2019 | Czech Republic, Greece, Italy, Lithuania, Norway, Poland, Romania, Russian Federation, Slovakia and Sweden | 2005-2006 | Randomized controlled trial | Community | 1887 | 12-22 months | Number of doses |
| Doshi et al, 2015 | Democratic Republic of Congo | 2010–2012 | Case-control | Case-Based (CB) measles surveillance system with laboratory confirmation | 2379 | 12-59 months | Malnutrition |
| Ferreira et al, 2018 | Brazil | 2007-2010 | Cohort study | Clinic | 121 | 12 months | Preterm birth |
| Ferreira et al, 2023 | Brazil | 2015-2016 | Cohort study | Community | 825 | 2 years | Malnutrition |
| Fouda et al, 2018 | Saudi Arabia | 2014-2017 | Cross-sectional | Clinic | 57 | 1-18 years | Treatment for comorbidities |
| Haralambieva et al, 2018 | USA | 2001-2011 | Cohort study | Community | 2872 | 11-40 years | Genetic variants |
| He at al, 2014 | China | NA | Randomized controlled trial | Community | 280 | 8, 12 months | Age at first dose |
| Jusko et al, 2019 | USA | 1999–2004 | Cross-sectional | Community | 7005 | 6-17 years | Toxins |
| Klein et al, 2021 | USA | 2018 | Randomized controlled trial | Community | 1736 | 12-15-month | Fever |
| Kontio et al, 2016 | Finland | 2013 | Cohort study | Clinic | 187 | 11-13, 17–19 months | Age at first dose |
| Martins et al, 2014 | Guinea-Bissau | 2003-2007 | Randomized controlled trial | Community+Hospital | 6417 | 4.5-36 months | Age at first dose |
| Martins et al, 2014 | Guinea-Bissau | 2003-2007 | Randomized controlled trial | Community | 900 | 4.5, 9, 18, and 24 months | Age at first dose |
| Mufson et al, 2015 | USA | 2009-2010 | Randomized controlled trial | Community | 1220 | 12-15 months | Vaccine strains |
| Pillsbury et al. 2015 | Australia | 2006-2012 | Case-control | National Notifiable Diseases Surveillance System (NNDSS) | 3987 | 1-15 years | Number of doses |
| Sánchez-Alemán et al, 2021 | Mexico | 2017 | Cross-sectional | Community | 416 | 6 to 13 years | Malnutrition |
| The MMR-158 Study Group, 2019 | USA | NA | Randomized controlled trial | Community | 3846 | 4-6 years | Vaccine strains |
| Zahraei et al, 2016 | Iran | 2013 | Cohort study | Urban Health Centers | 236 | 12 months | Malnutrition |
| Zimmermann et al, 2019 | Australia | 2013-2016 | Randomized controlled trial | Community | 471 | 7-13 months | Gender |
| Ferreira et al 2023 | Brazil | 2015-2016 | Cohort study | Urban Area | 1242 | 0-2 years | Malnutrition |
| Dhanorkar et al 2018 | India | 2018 | Cross-sectional study | Rural area | NA | - | Cold chain management |
| Doshi et al. 2015 | Democratic Republic of Congo (DRC). | 2010-2012 | Case Control Study | Community | 2379 | 12-59 months | Malnutrition |
| Geier et al 2019 | USA | 1990-2009 | Cohort study | Hospital | 101736 | 120 months | Age at first dose |
| ZAHRAEI ET AL. 2016 | Iran | 2013 | Cohort study | Urban Health Centers | 270 | 12-12 months 30 days | Malnutrition |
| Wang et al 2022 | China | 2017-2019 | Cohort study | Hospital | 110 | 0.8-14.4 years | Treatment for comorbidities |
| Pittet et al. 2024 | Switzerland | 2013-2022 | Cohort study | Hospital | 119 | 3.4-13.4 years | Treatment for comorbidities |
| Ovsyannikova et al. 2014 | USA | 2001-2007 | Cohort study | Community | 764 | 11-18 years | Genetic variants |
| Bianchi et al 2020 | Italy | 2014-2018 | Cohort study | Hospital | 4563 | All age groups | Age at first dose |
| Madi et al 2020 | Kuwait | 2017-2018 | Cross-sectional study | Hospital | 1000 | 5-20 years | Gender |
| Crom et al 2024 | USA | 2019-2022 | Cohort study | Hospital | 262 | 11-30 years | Treatment for comorbidities |
| Tomášková et al. 2018 | Czechia | 2013 | Cross-sectional study | Community | 3111 | 1-64 years | Age at first dose |
| Ghafoori et al 2024 | Iran | 2020 | Cohort study | Laboratory | 290 | 2-22 years | Gender |
| Musa et al. 2018 | Bosnia and Herzegovina | 2014-2015 | Cohort study | Community | 906 | 6-15 years | Number of doses |
| Koochakzadeh et al. 2014 | Iran | 2007-2008 | Case Control Study | Hospital | 90 | 2-16 years | Treatment for comorbidities |

Several factors were reported to likely influence VE including age (n=8); malnutrition (n=7); comorbidities (n=5); gender, number of doses (n=6); fever, genetic factors (n=2); as well as cold chain management, preterm birth, vaccine strain and toxin (n=1).

### Factors influencing VE

#### 1. Malnutrition

Child malnutrition is an important global public health issue with significant implications [21]. The relationship between malnutrition and childhood diseases like measles is particularly crucial, highlighting the need for measles vaccination for malnourished children [22].

A 2016 study conducted in Southeast Iran found a significantly lower seroconversion rate among malnourished children. The study involved 270 infants aged 12-months with 236 completing both pre- and post-vaccination blood sampling phases. After receiving the measles–mumps–rubella (MMR) vaccine, the seroconversion rate was 91.2% (95% confidence interval [CI]: 86.7-94.5) indicating a protective response against measles. However, stunting (height-for-age z-score <-2) was strongly associated with a lack of seroconversion (odds ratio=5.6; 95%CI: 1.7-18.2) [23].

Similar conclusions were drawn from a study in the Democratic Republic of Congo, which highlighted a strong association between malnourishment and measles cases. This study also looked at the malnourished children suffering from malaria infection. Lower VE was observed in malaria-endemic countries where many children suffer from malnutrition [24]. In Southern Mexico, low anti-measles serologic coverage (59%) was reported to be associated with age (lower antibodies in 10- to 13-year-old versus 6-year-old children), female sex, large family size (≥8 members), and underweight [25] Another study in Brazil found poverty and poor nutritional status to be closely associated with vaccine compliance and antibody positivity [26]. A study conducted in Eastern Mumbai slum areas in children with low nutritional status reported that VE was 64% (95%CI: 23-73%) and 70% (95%CI: 28-88%) for two doses among children <5 years and 5-15 years, respectively [27]. The above findings show a strong inverse relationship between malnutrition and VE.

#### 2. Age at first dose

Several studies examined whether the age of administration of the first dose impacts VE. In a district of Ethiopia, although the vaccination coverage rate was 95%, the overall measles VE was only 70.9% (95%CI: 65-79%) [28]. Only 45.1% of children received the first dose at 9 months and 35.3% received a second dose after 12 months. A study in China involving 280 infants showed that administering the MMR vaccine at 8 months, provides comparable immunogenicity as vaccination at 12 months [29]. Similarly, in Finland, where the MMR vaccination age was lowered to 12 months, seropositivity rates and IgG antibody concentrations in 187 3-year-old children were similar regardless of whether children were vaccinated at 11-13 months or 17-19 months [30]. In contrast, a retrospective longitudinal cohort study involving 101736 children reported that administering the MMR vaccine at 16-20 months of age was linked to higher VE compared to 12-15 months of age [31].

Additionally, in a retrospective cohort study conducted in Italy, a significant proportion of immunised individuals did not retain protective levels of anti-measles IgG antibodies 10 years later [32]. This proportion was higher among those vaccinated ≤15 months (20%) compared to those vaccinated at 16-23 months (17%) and ≥24 months (10%). In a study conducted in Guinea-Bissau, among 6417 children 77% had protective antibodies by 9 months after the first dose at 4.5 months. After a second dose at 9 months, 97% maintained antibody levels at 24 months. Among children who received MV at 9 months, 99% had antibody levels at 24 months even without a second dose at 18 months [33]. However, a cross-sectional study conducted in Pune, India, involving 600 children showed that despite receiving the MV at 9 months >25% of children became susceptible by 12-15 months [34].

#### 3. Vaccine strains

MVs are live attenuated and derived from various strains, with globally used strains including Edmonston-Zagreb, Schwarz, Moraten, Leningrad-16, Hu-191 (Shanghai- 191), CAM-70, TD-97 [35]. In India, the Edmonston-Zagreb and Schwarz strains are primarily used [36, 37]. The Hu-191 strain in the MMR vaccine achieved a 100% seropositivity rate for measles when the first dose was administered at 8 or 12 months [29], while the Edmonston-Zagreb strain reached up to 97% seropositivity for early vaccination at ≤6 months and 99% for the first dose at 9 months [33]. Mufson et al[38] evaluated the immunogenicity and safety of various MMR vaccine strains like Priorix™ (MMR-RIT) and MMRII. In a study of 1220 healthy children aged 12–15 months, results showed high seroresponse rates for both MMR-RIT (98.3-99.2%) and MMRII (99.6%) strains. Similarly, in an RCT[39] comparable immune responses was observed for the MMR-RIT (Priorix, GSK) and MMR II (M-M-R II, Merck & Co Inc.) vaccines when administered to 4011 children aged 4-6 years as a second dose, either alone or in combination with the DTaP-IPV and varicella vaccines. These findings suggest that the effect on VE is comparable between vaccine strains.

#### 4. Fever

Fever is a common side effect following measles vaccination. Predictably, measles vaccination may induce fever in 5%-15% of vaccine recipients 7-10 days post-vaccination [40, 41]. Fever after vaccination may indicate an innate and/or cell-mediated immune response that precedes the humoral response [42].

A study found that the geometric mean titres (GMT) against measles in children without and with fever after MMR vaccination were 2918 (95%CI: 2318-3673) and 4609 (95%CI: 3629-5853), respectively [43]. These results suggest that fever following vaccination is strongly linked to a higher immune response.

#### 5. Cold chain management

Cold-chain management is crucial for maintaining VE, particularly for temperature-sensitive vaccines like MV. While stable between -70°C and -20°C, the MV rapidly loses potency once reconstituted and exposed to higher temperatures, losing about 50% at 20°C in one hour and nearly all at 37°C [28]. Proper cold-chain management requires keeping vaccines at −20°C in national stores and 2-8°C at health facilities, with usage within 4 hours post-reconstitution [44, 45]. Failure to maintain these conditions can reduce VE, increasing the risk of outbreaks in areas with poor infrastructure [28].

#### 6. Number of doses

Two doses of MV are recommended to ensure robust immunity and prevent outbreaks, as not all children develop immunity from the first dose [3]. According to the Center for Disease Control and Prevention (CDC), a single dose of MV is approximately 93% effective at preventing measles if exposed to the virus, while two doses are approximately 97% effective [46]. An RCT conducted across several European countries examined the long-term effects of 1 and 2 doses of MMR- containing vaccines administered in the second year of life [47]. The study found that these vaccines induced antibody responses that persisted post-vaccination with high seropositivity rates even a decade later regardless of the vaccine administered and schedule. A cohort study conducted in the Federation of Bosnia and Herzegovina found that the effectiveness of MV was 91.9% (95%CI: 81.4–96.4%) for a single dose and 97.3% (95%CI: 95.5–98.4%) for two doses [48]. Another study in Australia assessed the effectiveness of MV at the population level using national notification data from 2006-2012 [49]. Analysing cases of measles in children born after 1996, the study found the estimated effectiveness of MV to be 96.7% (95%CI: 94.5–98.0%) and 99.7% (95%CI: 99.2–99.9%) for one and two doses, respectively. Overall, VE for ≥1 dose was 98.7% (95%CI: 97.9-99.2%) suggesting that two doses are more effective than one dose of MV.

#### 7. Preterm birth

A prospective study compared the immune response to MV between infants born prematurely and those born at full term [50]. The study included 65 premature infants (birth weight <1500 g) and 56 full-term infants aged 12 months. The results showed that both groups had similar rates of immunity following vaccination (antibody levels: 2.393 vs. 2.412 UI/mL; *p*=0.970). Overall, the study concluded that humoral responses to measles did not vary among premature and full-term infants.

#### 8. Toxins

A cross-sectional study in the US analysed the link between blood lead levels and antibody responses to measles, mumps, and rubella in 7005 children aged 6-17 years, using data from the 1999-2004 National Health and Nutrition Examination Survey [51]. Children with blood lead levels of 1-5 µg/dL had 11% (95%CI: -16, -5) lower anti-measles antibodies and were twice as likely to be seronegative for measles. This suggests that lead levels even below the CDC’s 5 µg/dL action level may impair immune function and VE.

#### 9. Genetic variants

A genome-wide association study was conducted in the USA focusing on measles-specific neutralising antibodies and IFNγ ELISPOT responses in 2872 subjects [52]. The findings revealed that common single-nucleotide polymorphisms (SNP) in the CD46 and IFI44L genes are associated with measles-specific humoral immunity.

This study underscores the significance of studying the relationship of genetic variants to the inter-individual variation in immune response following live measles vaccination. Furthermore, a cohort study with 764 healthy children aged 11-18 (Caucasian-American, 80.6%; African-American, 11.6%) from Rochester, was conducted to measure the immune response to MV[53]. Post-vaccination immune responses was found to be associated with genetic polymorphism in the DDX58 gene (rs669260). Overall, variation in genes may lead to differences in VE.

#### 10. Treatment for comorbidities

Children with acute lymphoblastic leukaemia (ALL) face a risk of infection from viral diseases like measles after completing chemotherapy, which could be mitigated by MMR vaccination. Results of a retrospective cross-sectional study in Saudi Arabia showed that of 57 children who survived ALL, 35 (61.4%) were seropositive, while 22 (38.6%) were seronegative [54]. Notably, children <5 years had higher rates of seronegativity and protection decreased with time since ALL treatment. Booster vaccination resulted in a seroconversion of 57.1%. Similarly, in another study in the USA, among the 262 leukaemia, solid tumour, or brain tumour survivors, 110 (42%) tested negative for anti-measles IgG antibodies, while 152 (58%) tested positive [55].

A study involving 90 patients treated for ALL in Iran showed that seropositivity to measles was achieved if MMR was administered ≥3 months after chemotherapy [56]. Similarly, a study in China analysed clinical data from 110 children who completed chemotherapy and were subsequently revaccinated with the MMR vaccine[57]. Overall, 70 children were seronegative after chemotherapy. The mean measles antibody titres were 254.4±67.46 mIU/mL at baseline and increased to 1471.0±210.0 mIU/mL and 1872.0±262.1 mIU/mL at 1- and 6-months post-vaccination, but decreased to 1710.0±238.6 mIU/mL at 12 months.

In a cohort study, 19 liver transplant recipients were vaccinated based on the seroprotection status and were followed up to 9 years post vaccination[58]. During the follow-up period, approximately half of the recipients lost their seroprotection likely due to biliary atresia, treatment with anti-rejection drugs, or MMR vaccination before transplantation. These studies highlight the need for periodic monitoring of antibody titres in children with immunocompromising conditions and the potential requirement for booster doses to maintain adequate antibody levels.

#### 11. Gender

VE may vary between males and females [30, 59–61]. A study using samples from Iran’s National Measles Laboratory found that the geometric mean anti-measles IgG antibody level was higher in females, at 554.9 mIU/mL, compared to 468.4 mIU/mL in males [59]. Madi et al conducted an age-stratified serological study in Kuwait with 1000 participants. [60]. In that study, females had a higher GMT of 5.5 IU/ml (95%CI: 5.9-5.0) compared to 4.7 IU/ml (95%CI: 5.2-4.3) in males. Similar observations were reported in other studies as well [30, 61]. Boys vaccinated at 11-13 months in Finland, had lower antibody concentrations compared to girls and increasing the age of vaccination appeared to improve antibody responses in boys [30]. This indicates that females may have a stronger immune response to the MV than males.

### Risk of bias within studies

Risk of bias was low (80-100%) for most of the included studies. Only three cohort studies and one cross-sectional study showed medium risk of bias (50-79%). No other risks were identified and no studies were excluded based on this assessment.

### Modelling the impact of malnutrition on VE

As discussed earlier, VE is implicitly influenced by the proportion of malnourished children under 5 years of age. Due to a limitation of data published after 2014, we utilised data before 2014 to explore this relationship. We based our analysis on VE data available for 5 regions in India, namely Kangra (Himachal Pradesh, 2006), Purulia (West Bengal; 2005, 2006), Surat (Gujarat, 2003) and Cuddalore (Tamil Nadu, 2004) from various cohort studies [62–64]. The proportion of severely malnourished children (below -2 SD) of the above states is obtained from the National Family Health Survey (NFHS) 5.

To understand the relationship between malnutrition and VE, we fit a simple linear regression model to the data. In **Figure 2** we present the fit of two linear models that we have implemented along with 95% CIs. The former corresponds to a standard simple linear regression model and later is a model where the intercept parameter is constrained to not exceed the baseline VE which we considered to be 98%. We observe a decreasing trend in VE with an increase in undernourished percentage in both the model setups, but they vary in terms of magnitude of the slope parameter. For Model-1 the estimated coefficients of intercept and slope are (150.866, -2.036) and for the constrained model (Model-2) estimates are (98, -0.6541). The observed findings are statistically significant at 10% level of significance, suggesting moderate evidence of an inverse relationship between VE and undernourishment. However, further investigation with additional data is recommended to strengthen this result.

**Figure 2.**
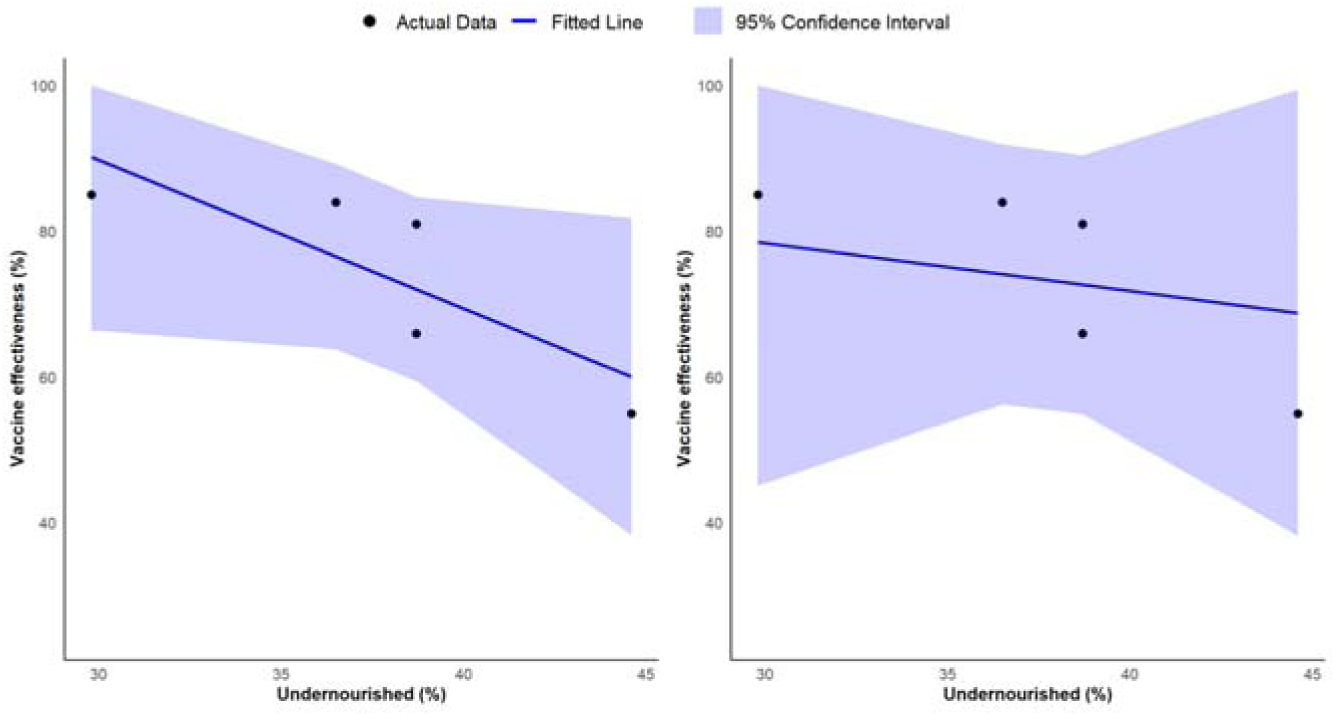
Relationship between vaccine effectiveness and percentage of undernourished children. Model fit and 95% confidence intervals for standard linear model (Model-1; left) and constrained linear model (Model-2; right).

Furthermore, we use the results of Model-2 to estimate a possible decrease in VE due to undernutrition for all Indian states for a given proportion of undernourished children from (NFHS-5). The expected percentage decrease in VE due to malnutrition is calculated by multiplying the observed percentage of undernourished individuals by 0.6541, the slope estimate derived from Model 2. The results thus obtained for all the states and union territories of India are presented in **Figure 3**.

**Figure 3.**
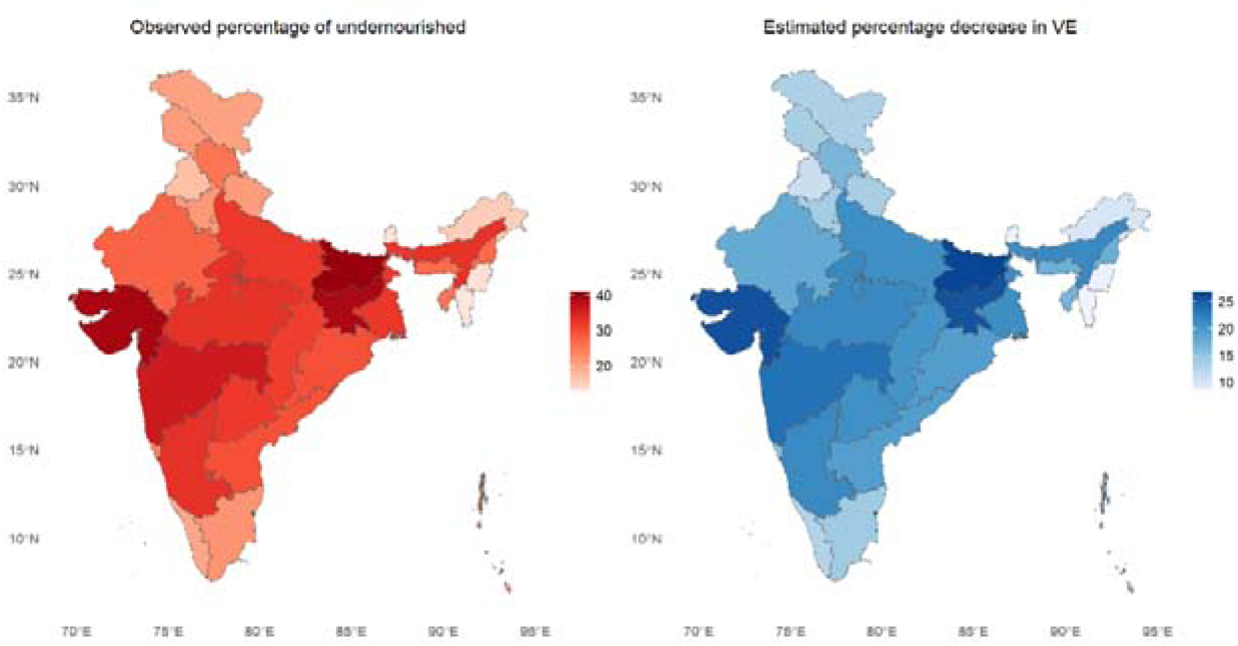
Spatial distribution of percentage of undernourished children (left) and model estimated percentage decrease in VE due to undernourishment (right). E, east; N, north; VE, vaccine effectiveness [Shape file source: https://onlinemaps.surveyofindia.gov.in; Software: R]

### Impact of VE on measles incidence

Although high vaccination coverage is essential, VE also plays a critical role in determining the extent of an epidemic outbreak. For illustration purposes, we use an SI-type epidemic model [19, 20] (details of the model are provided in the supplementary material) with an initial population size of 5 X 105, the estimates of α=0.97 and birth rate of 0.015/26 per bi-weekly period. The seasonal measles transmission rate β is taken from previous studies [65, 66]. Vaccine coverages are assumed to be constant over time, with (i) high coverage: 90% for the first dose and 85% for the second dose; (ii) medium coverage: 85% for the first dose and 75% for the second dose.

Some modelling studies assume high average VE in the range of 90-99% [20, 67]. However, VE may be significantly lower in certain regions. For instance, in South East Asian regions, the VE is 77% [4, 27]. Our analysis compares a baseline scenario without vaccination to two different vaccination scenarios with average VE of 95% and 77%, of two doses respectively. **Figure 4** displays the cases averted over a span of 20 years. The findings indicate that while vaccination significantly reduces cumulative cases compared to the no-vaccination baseline, the effectiveness of the vaccine is also crucial. Specifically, for high coverage, a vaccine with 95% effectiveness can avert approximately 94.54% of cases, whereas a vaccine with 77% effectiveness can avert about 82.00% (black, **Figure 4**). Similarly, for medium coverage, a vaccine with 95% effectiveness can avert 91.07% of cases, whereas a vaccine with 77% effectiveness can avert about 77.83% (grey bars, **Figure 4**). This underscores the importance of VE in managing disease outbreaks and achieving global measles elimination targets.

**Figure 4.**
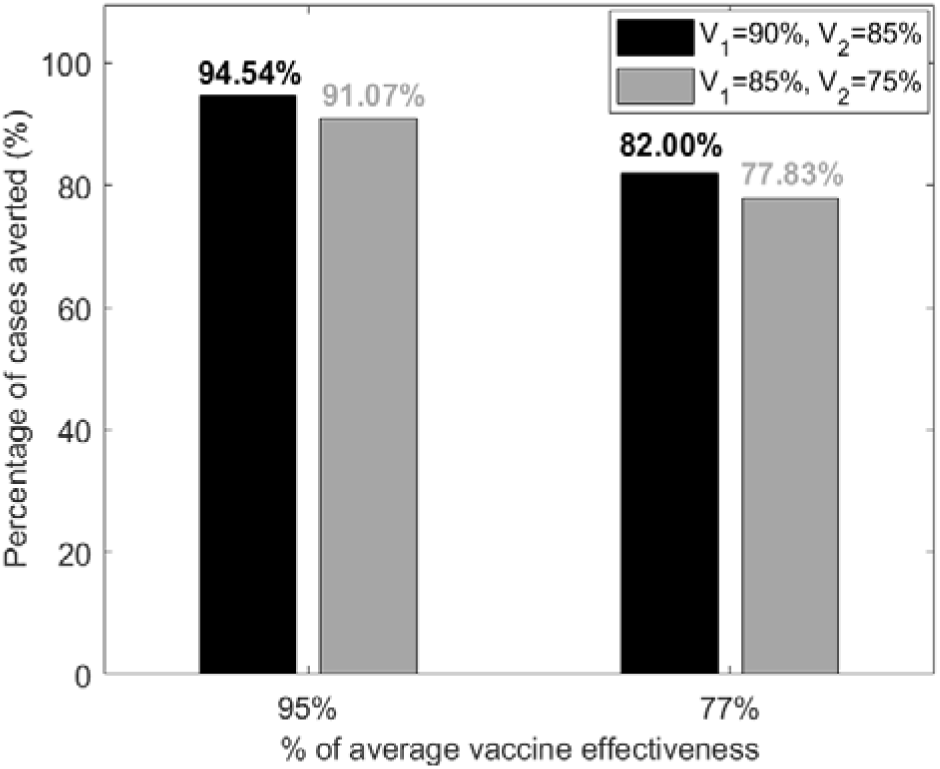
Percentage of cases averted under two different vaccination coverage scenarios. High coverage (V₁ = 90%, V₂ = 85%) is represented by black bars, while medium coverage (V₁ = 85%, V₂ = 75%) is depicted by grey bars. The two bars on the left show results for 95% VE, and the two bars on the right represent outcomes with 77% VE. V1 and V2 refer to first and second doses, respectively.

## DISCUSSION

This systematic review highlights several factors that may contribute to the reduced effectiveness of MV, which could eventually lead to outbreaks of measles reported in various parts of the world. An important factor to be considered in low-and-middle-income countries (LMIC) is malnutrition among children. Several studies reported that malnutrition was associated with low seroconversion rates in children receiving MV [23–27]. Similar observations have been reported for other vaccine-preventable diseases (VPD) such as rotavirus [68] and tetanus [69]. Malnourished children are at a greater risk of infections [70] and contribute to nearly half of the deaths in children <5 years of age globally [71]. In India, the mean prevalence of malnourished children was 7.56% between 2019-2021 [72]. Therefore, vaccination strategies in this population need to be considered carefully.

Furthermore, from our model fit, we noted that higher malnutrition could be a potential factor to lower VE. Also, variation in the malnutrition status at the country or state level may cause an associated variation in the VE. The simulation results of the SI epidemiological model illustrate that not only vaccine coverage but also VE plays a major part in disease transmission and future outbreaks. Modelling scenarios with higher VE may result in a more optimistic/misleading measles incidence scenario compared to lower VE. However, because actual VE may not be high across regions, region-specific VE should be considered to estimate measles incidence more accurately. Overestimating VE could also adversely impact the measles elimination deadlines.

Several other factors also have an effect on VE. Consistent with studies in other VPDs [15], genetics can play a role in determining the immune response to MV [52, 53]. The common SNPs in the CD46 and IFI44L genes and the DDX58 gene are associated with MV-specific immunity. Treatment for comorbidities such as cancer was also reported to impact VE. These studies report that chemotherapy resulted in seronegativity in vaccinated children [54, 57]. Therefore, booster doses or vaccination after chemotherapy has been recommended. A previous review has outlined the effects of other cancer therapies on vaccination and has provided points to consider for patients receiving vaccination [73]. Consistent with previous findings for other vaccines [15], several studies included in this analysis report that the immune response to MV is stronger in females than males outlining the role of gender in VE [30, 59–61]. In addition to patient-related factors, the storage and distribution of vaccines or cold-chain management also impacts measles VE with higher temperatures reducing vaccine potency.

The effect of some factors identified in our analysis including age, vaccine strain, and preterm birth on VE were either inconclusive or did not show an effect. Studies on the impact of age reported conflicting results with some studies suggesting comparable protection for two different age groups [29, 30] and different VE for vaccination at 9 months [33, 34], while others reported better or long-term VE when infants were vaccinated at 16-24 months versus <15 months [31, 32]. Different vaccine strains showed comparable effects on VE. Other studies showed that VE did not vary between children born preterm or full-term. Only limited studies were obtained for other factors such as the effect of toxins and were insufficient to draw a conclusion on their impact on VE.

Our study had few limitations. Due to a limited number of studies and available data as well as heterogeneity among the studies, a meta-analysis was not feasible.

Additionally, since very limited published data on VE was available from India, we carried out a regression analysis using data before 2014. The data on VE at the regional level in India and survey data on undernourished children provided a foundation for our analysis. As more data on VE and other potential factors become available, the application of advanced statistical tools will enable a deeper and more precise understanding of how these factors influence VE leading to more informed insights and effective interventions.

Overall, this analysis outlines some potential factors that may impact VE thereby laying the foundation for future work. One of the proposed areas to explore is to address malnutrition through nutritional supplementation programs and micronutrient supplementation, such as vitamin A, to strengthen the immune system of children in LMICs. In this context, various scenario analyses have been conducted to assess the impact of such nutritional programs in the reduction of measles incidence [74].

Furthermore, given the limited region-specific studies currently available on the impact of genetic factors on measles, additional studies in LMIC’s are warranted. Moreover, from a modelling perspective, to better capture the dynamics of measles transmission, we plan to model VE as a function of the potential factors discussed in this work rather than being treated as constant.

In conclusion, understanding the underlying causes of variation in the VE and identifying geographical regions with lower VE is crucial for measles control and elimination. Thus, policymakers while deciding disease control measures and setting elimination targets will need to have a clear understanding of the variation in VE and the factors causing this variation.

## AUTHOR CONTRIBUTIONS

SG, DK: Conceptualization, data curation, screening, analysis, epidemiological modelling, and writing. NM, Conceptualization, data curation, screening, analysis, and writing. SNS: Conceptualization, data curation, screening, analysis, writing, editing and formatting. ADR: Reviewing. SM: Conceptualization, supervising, writing and reviewing. All authors read and approved the final draft of the manuscript.

## Data Availability

All data produced in the present work are taken from published research works and are contained in the manuscript.

