## Supplementary file for "A systematic review and modelling insights of factors impacting measles vaccine effectiveness"

| **Supplementary Table I. Quality assessment of case-control studies** | | | | | | | | | | | **Score**  **(%)** | **Risk** |
| --- | --- | --- | --- | --- | --- | --- | --- | --- | --- | --- | --- | --- |
| **Author name and year** | **Q1** | **Q2** | **Q3** | **Q4** | **Q5** | **Q6** | **Q7** | **Q8** | **Q9** | **Q10** |  |  |
| Bati et al, 2021 | Y | N | Y | Y | Y | Y | Y | Y | Y | Y | 90 | Low |
| Pillsbury et al, 2015 | Y | Y | Y | Y | Y | Y | Y | Y | Y | Y | 100 | Low |
| Doshi et al, 2015 | Y | Y | Y | Y | Y | Y | Y | Y | Y | Y | 100 | Low |
| Hi et al, 2015 | Y | Y | Y | Y | Y | N | N | Y | Y | Y | 80 | Low |
| Koochakzadeh et al, 2014 | Y | Y | Y | Y | Y | Y | Y | Y | Y | Y | 100 | Low |

Q1. Were the groups comparable other than the presence of disease in cases or the absence of disease in controls?

Q2. Were cases and controls matched appropriately?

Q3. Were the same criteria used for identification of cases and controls?

Q4. Was exposure measured in a standard, valid and reliable way?

Q5. Was exposure measured in the same way for cases and controls?

Q6. Were confounding factors identified?

Q7. Were strategies to deal with confounding factors stated?

Q8. Were outcomes assessed in a standard, valid and reliable way for cases and controls?

Q9. Was the exposure period of interest long enough to be meaningful?

Q10. Was appropriate statistical analysis used?

| **Supplementary Table II. Quality assessment of randomized controlled trials** | | | | | | | | | | | | | | **Score**  **(%)** | **Risk** |
| --- | --- | --- | --- | --- | --- | --- | --- | --- | --- | --- | --- | --- | --- | --- | --- |
| **Author name and year** | **Q1** | **Q2** | **Q3** | **Q4** | **Q5** | **Q6** | **Q7** | **Q8** | **Q9** | **Q10** | **Q11** | **Q12** | **Q13** |  |  |
| He at al, 2014 | Y | Y | Y | N | N | Y | Y | Y | Y | Y | Y | Y | Y | 84 | Low |
| Martins et al, 2014 | Y | Y | Y | U | U | Y | Y | Y | Y | Y | Y | Y | Y | 84 | Low |
| Martins et al, 2014 | Y | Y | Y | U | U | Y | Y | Y | Y | Y | Y | Y | Y | 84 | Low |
| Carryn et al, 2019 | Y | Y | Y | Y | Y | Y | Y | Y | Y | Y | Y | Y | Y | 100 | Low |
| The MMR-158 Study Group, 2019 | Y | Y | Y | Y | Y | Y | Y | Y | Y | Y | Y | Y | Y | 100 | Low |
| Mufson et al, 2015 | Y | Y | Y | Y | Y | Y | Y | Y | Y | Y | Y | Y | Y | 100 | Low |
| Zimmermann et al, 2019 | Y | Y | Y | Y | Y | Y | Y | Y | Y | Y | Y | Y | Y | 100 | Low |
| Klein et al, 2021 | Y | Y | Y | Y | Y | Y | Y | Y | Y | U | Y | N | Y | 84 | Low |

Question 1: Was true randomization used for assignment of participants to treatment groups?

Question 2: Was allocation to groups concealed?

Question 3: Were treatment groups similar at the baseline?

Question 4: Were participants blind to treatment assignment?

Question 5: Were those delivering the treatment blind to treatment assignment?

Question 6: Were treatment groups treated identically other than the intervention of interest?

Question 7: Were outcome assessors blind to treatment assignment?

Question 8: Were outcomes measured in the same way for treatment groups?

Question 9: Were outcomes measured in a reliable way?

Question 10: Was follow up complete and if not, were differences between groups in terms of their follow up adequately described and analysed?

Question 11: Were participants analysed in the groups to which they were randomized?

Question 12: Was appropriate statistical analysis used?

Question 13: Was the trial design appropriate and any deviations from the standard RCT design (individual randomization, parallel groups) accounted for in the conduct and analysis of the trial?

| **Supplementary Table III. Quality assessment of cohort studies** | | | | | | | | | | | | **Score**  **(%)** | **Risk** |
| --- | --- | --- | --- | --- | --- | --- | --- | --- | --- | --- | --- | --- | --- |
| **Author name and year** | **Q1** | **Q2** | **Q3** | **Q4** | **Q5** | **Q6** | **Q7** | **Q8** | **Q9** | **Q10** | **Q11** |  |  |
| Kontio et al, 2016 | Y | Y | Y | Y | Y | Y | Y | Y | Y | Y | Y | 100 | Low |
| Haralambieva et al, 2018 | Y | Y | Y | Y | Y | Y | Y | Y | Y | Y | Y | 100 | Low |
| Ferreira et al, 2018 | Y | Y | Y | Y | Y | Y | Y | Y | Y | Y | Y | 100 | Low |
| Zahraei et al, 2016 | Y | Y | Y | Y | Y | Y | Y | Y | Y | Y | Y | 100 | Low |
| Ferreira et al, 2023 | Y | Y | Y | Y | Y | Y | Y | Y | Y | Y | Y | 100 | Low |
| Ferreira et al, 2023 | U | U | Y | Y | Y | Y | Y | Y | Y | U | Y | 73 | Medium |
| Zahraei et al, 2016 | Y | Y | Y | Y | Y | Y | Y | Y | Y | Y | Y | 100 | Low |
| Bianchi et al, 2020 | Y | Y | Y | Y | Y | Y | Y | Y | Y | Y | Y | 100 | Low |
| Crom et al, 2024 | Y | Y | Y | Y | Y | Y | Y | Y | Y | N | Y | 90 | Low |
| Ghafoori et al, 2024 | Y | Y | Y | Y | Y | Y | Y | Y | Y | N | Y | 90 | Low |
| Musa et al, 2018 | Y | Y | Y | N | N | Y | Y | Y | Y | N | Y | 73 | Medium |
| Ovsyannikova et al, 2014 | Y | Y | Y | Y | Y | Y | Y | U | U | U | Y | 73 | Medium |
| Pittet et al, 2024 | Y | Y | Y | Y | Y | Y | Y | Y | Y | Y | Y | 100 | Low |
| Wanget al, 2023 | Y | Y | Y | Y | Y | Y | Y | Y | Y | Y | Y | 100 | Low |
| Geier et al, 2019 | Y | Y | Y | Y | Y | Y | Y | Y | Y | Y | Y | 100 | Low |

1. Were the two groups similar and recruited from the same population?

2. Were the exposures measured similarly to assign people to both exposed and unexposed groups?

3. Was the exposure measured in a valid and reliable way?

4. Were confounding factors identified?

5. Were strategies to deal with confounding factors stated?

6. Were the groups/participants free of the outcome at the start of the study (or at the moment of exposure)?

7. Were the outcomes measured in a valid and reliable way?

8. Was the follow up time reported and sufficient to be long enough for outcomes to occur?

9. Was follow up complete, and if not, were the reasons to loss to follow up described and explored?

10. Were strategies to address incomplete follow up utilized?

11. Was appropriate statistical analysis used?

| **Supplementary Table IV. Quality assessment of cross-sectional studies** | | | | | | | | | **Score**  **(%)** | **Risk** |
| --- | --- | --- | --- | --- | --- | --- | --- | --- | --- | --- |
| **Author name and year** | **Q1** | **Q2** | **Q3** | **Q4** | **Q5** | **Q6** | **Q7** | **Q8** |  |  |
| Jusko et al, 2019 | Y | Y | Y | Y | Y | Y | Y | Y | 100 | Low |
| Fouda et al, 2018 | Y | Y | Y | Y | Y | Y | Y | Y | 100 | Low |
| Sánchez-Alemán et al, 2021 | Y | Y | Y | Y | Y | Y | Y | Y | 100 | Low |
| Tomášková et al, 2018 | Y | Y | Y | Y | Y | Y | Y | Y | 100 | Low |
| Madi et al, 2020 | Y | Y | Y | Y | Y | Y | Y | Y | 100 | Low |
| Dhanorkar et al, 2018 | Y | Y | Y | Y | U | U | Y | N | 75 | Medium |

1. Were the criteria for inclusion in the sample clearly defined?

2. Were the study subjects and the setting described in detail?

3. Was the exposure measured in a valid and reliable way?

4. Were objective, standard criteria used for measurement of the condition?

5. Were confounding factors identified?

6. Were strategies to deal with confounding factors stated?

7. Were the outcomes measured in a valid and reliable way?

8. Was appropriate statistical analysis used?

**Model description**

$$S_{t}=B_{t}+S_{t-1}-I_{t} ,$$

$$I_{t}=\beta_{t}{I^{\alpha}}_{t-1}S_{t-1} ,$$

where, $\beta_{t}$ is the disease transmission rate which takes into account the seasonality, $\alpha$ is the mixing parameter, $S_{t}$ and $I_{t}$ denote the susceptible and infected individuals at time $t$. $B_{t}$ is the adjusted birth rate at time $t$given by:

$B_{t}=\hat{B_{t}} (1-\epsilon_{1}V_{1}(1-V_{2})-\epsilon_{2}V_{1}V_{2}$),

where, $\hat{B_{t}}$ is the new births at time $t,$ $\epsilon_{1}$and $\epsilon_{2}$are the VE after the first and second dose of MV, respectively. The coverages of the first and second doses are denoted as $V_{1}$ and $V_{2}$, respectively.
